# Mapping COVID-19 vaccine acceptance and uptake amongst Chinese residents: A systematic review and meta-analysis

**DOI:** 10.1101/2023.08.09.23293915

**Authors:** Hassan Masood, Syed Irfan

**Affiliations:** Faculty of Health Sciences, McMaster University, Hamilton, Canada

## Abstract

**Objective:** Controlling the COVID-19 pandemic depends on the widespread acceptance of vaccination. Vaccine hesitancy is a growing area of concern in China. The aim of the study is to map the overall acceptance and uptake rates of COVID-19 vaccines across different groups.

**Methods:** Five peer-reviewed databases bases were searched (PubMed, EMBASE, Web of Science, EBSCO, and Scopus). Studies that conducted cross-sectional surveys in China to understand the acceptance/willingness to receive COVID-19 vaccines were included.

**Results:** Among 2420 identified studies, 47 studies with 327,046 participants were eligible for data extraction. Males had a higher uptake of COVID-19 vaccines (OR=1.17; 95% CI:1.08 - 1.27) along with Chinese residents with ≥ 5000 RMB monthly income (OR=1.08; 95% CI:1.02 - 1.14).

**Conclusion:** COVID-19 vaccination uptake rates in China need to be improved. To inform public health decisions, continuous vaccination uptake monitoring is required.

## 1. Introduction

### 1.1. Background of COVID-19

The emergence of coronavirus disease 2019 (COVID-19), a respiratory illness caused by the virus SARS-CoV-2, has resulted in a global pandemic since its breakout in 2019 [1]. More than 213 counties have reported COVID-19 outbreaks [2]. As of November 2022, there were more than 600 million confirmed cases of COVID-19 across the globe, resulting in approximately 7 million mortalities [1]. Looking at China alone, the World Health Organization (WHO) reported over 99 million COVID-19 cases and 121,000 deaths [3]. COVID-19 continues to impact public health as novel contagious variants of SARS-CoV-2 can affect fully immunized individuals [4]. According to data published by the WHO, different population groups are impacted disproportionately [4]. For instance, healthcare workers are at higher risk of contracting COVID-19 infection due to their occupational exposure to patients [4]. COVID-19 cases among young adults have a higher probability of resulting in asymptomatic infections compared to their older counterparts [1]. Similarly, sociodemographic factors such as age, gender, and level of knowledge influence the rate of uptake of vaccines and contribute to vaccine hesitancy [4] Scientific literature also indicates that the COVID-19 pandemic has disrupted the functionality of different systems, such as health and education [1]. In addition to the virus’ physical health impacts, reports show that there are also severe mental health repercussions of this deadly COVID-19 infection [1].

### 1.2. Importance of vaccines

The cataclysmic consequences of the COVID-19 pandemic have resulted in rapid vaccine research and development [5]. Vaccine development is a rigorous and time-consuming process, and it can take up to 10 years to develop a safe and effective vaccine [5]. However, the deadly impacts of COVID-19 have increased the pace of vaccine production, where vaccines were developed within one year [5]. Scientific evidence emphasizes the importance of vaccines as they are critical to ensure protection amongst the public [5]. Promoting the uptake of vaccination services results in the state of herd immunity, which is a fundamental method to control the spread of such infections [2] Considering the delta variant of the SARS-CoV-2, roughly 85% of the population should gain immunity to COVID-19 either via natural infection or by uptaking vaccination services [5]. However, the omicron variant of SARS-CoV-2 displayed stronger abilities to fight neutralizing antibodies produced by vaccines [5]. In this case, scientific evidence suggests that more than 85% of the population needs to be immunized to achieve herd immunity [5]. Therefore, achieving high vaccine coverage within a population is key to reducing the severity of the COVID-19 pandemic [2].

### 1.3. COVID-19 Protocols in China

In order to fight the COVID-19 pandemic, the government of China established several safety protocols [2]. These restrictions included lockdown, isolation, quarantine mandates, travel restrictions and prohibition of mass gatherings [2]. While these protocols displayed increased success over time in controlling the spread of the infection, they yielded negative consequences for economic growth and social development [2]. Therefore, several restrictions were lifted in China, and the focus shifted more toward vaccine development and promotion amongst the general public [2].

#### 1.3.1. China’s Zero-covid Policy

China makes a unique global case due to its strict zero Covid policy implementation over three years. When the COVID-19 pandemic was declared, the Chinese government viewed it as a major threat and placed strict restrictions to keep the COVID-19 cases close to zero [6]. These steps were collectively termed China’s COVID zero policy [6]. While these strategies effectively controlled the spread, they severely affected the economy and fueled protests across many cities [7]. Government finances were strained as most of the funds were directed toward COVID-19 safety protocols [7].

#### 1.3.2. Overnight wholesale re-opening

Due to these substantial economic and social losses, the Chinese government decided to ease the stringent regime. In December 2022, China lifted the restrictions to open up the factories, companies, stores and restaurants in an attempt to return back to pre-pandemic conditions [8]. The cases surged dramatically, and an extremely low vaccination rate was reported among older adults [9]. After relaxing the restrictions, over a million COVID-19 cases and 5000 deaths per day have been reported [9].

This recent surge of COVID-19 cases has severely impacted two populations: the elderly and those with underlying medical conditions. A study conducted by Ioannidis et al. [10] explored the increase in cases and deaths after the abandonment of the zero COVID policy and found that from December 2023 till the summer of 2023, 691,219 new cases of COVID-19 were recorded in China [10]. The majority of the fatalities were observed in senior citizens that were aged 60 years and above. In Hong Kong alone, 70.6% of deaths due to COVID-19 were among those aged 80 and above [10] The second highest deaths were recorded for those that were 70-79 years old (Ioannidis et al., 2023). These disproportionate fatal outcomes among the elderly are of high concern considering that as of December 2022, only 40% of the people over the age of 80 are vaccinated against COVID-19 [10]. This figure is even worse in some parts of China such as Hong Kong, where only 25% of people over the age of 80 are vaccinated [10].

### 1.4. Vaccine Development in China

After extensive research and testing, the China National Medical Products Administration (CNMPA) approved two COVID-19 vaccines: Sinopharm and Sinovac [11]. Sinopharm was approved in December 2020, whereas Sinovac was approved in February 2021 [11]. According to recent studies, there are three types of vaccines that are available in China for uptake [1]. These include the single-dose adenovirus vector vaccine, the 2-dose inactivated vaccine and the 3-dose recombinant subunit vaccine [1]. Most vaccine research in China was conducted before the approval of Chinese vaccines, whereby the population’s perceptions were based on the benefits and barriers to hypothetical COVID-19 vaccines [11]. Findings by Lin et al. [12] show that some Chinese residents prefer domestic vaccines over imported ones. Therefore, it is crucial to conduct in-depth research that aims to evaluate the different COVID-19 vaccine uptake rates along with reasons to create targeted approaches.

### 1.5. Vaccine Hesitancy in China

Due to widespread misinformation, public vaccine acceptance can vary over time. Social media is one of the avenues where misinformation is spread [13]. “Weibo” is China’s most popular social media platform, with reportedly 500 million active users [13]. Since the rise of the COVID-19 pandemic, Weibo has been one of the outlets where the general public in China has been having discussions regarding COVID-19 vaccines [13]. Due to the vast outreach of Wiebo, it is cited as one of the sources that breed COVID-19 vaccine hesitancy amongst the Chinese population in particular [13].

Research conducted by Liu et al. [14] demonstrated that there is variation in vaccine hesitancy between China and the United States (US). Chinese residents hold a different viewpoint, as they reported being more concerned about the adverse effects of COVID-19 vaccines (19.68%) compared to US residents (6.12%) [14]. While the overall prevalence of hesitancy was found to be high in both countries, the reasons for vaccine refusal were drastically different. In order to develop innovative and personalized solutions to achieving herd immunity, it is important to understand the unique challenges that are at play within different countries.

While significant research has examined the acceptance of COVID-19 vaccines, there are few studies that have systematically reviewed and compiled the current evidence [5]. As China is a heavily populated country with complex national conditions, it is imperative to investigate the COVID-19 vaccine acceptance, uptake, and reasons for vaccine hesitancy. By understanding the factors fueling vaccine refusal, tailored strategies can be undertaken to increase the efficacy of COVID-19 vaccination campaigns in China.

### 1.6. Research objective

This systematic review and meta-analysis aims to examine the 1) acceptance and uptake of COVID-19 vaccines across different population groups (adults, healthcare workers, patients with chronic diseases, pregnant women, university students, and parents) in China; 2) compare the differences in uptake rates across diverse subgroups based on their sociodemographic characteristics; and 3) common reasons for vaccine refusal. The population subgroups investigated in this study include age, gender, income, and level of education. This in-depth analysis will add context to current research and help guide vaccine promotion strategies in China by understanding each subgroup’s concerns, beliefs, and needs. The findings can also function as a reference for future studies that investigate COVID-19 vaccination beliefs in China.

## 2. Materials and methods

### 2.1. Data Sources and Strategy

This systematic review and meta-analysis was developed according to the Preferred Reporting Items for Systematic Reviews and Meta-Analyses (PRISMA) guidelines [15]. The following peer-reviewed databases were searched: PubMed, EMBASE, Web of Science, EBSCO, and Scopus.

Within these databases, the following search terms were employed: coronavirus terms (“coronavirus disease”[All Fields] OR “coronavirus”[MeSH Terms] OR “coronavirus”[All Fields] OR “coronaviruses”[All Fields] OR “2019-nCoV”[All Fields] OR “2019ncov”[All Fields] OR “covid 19”[All Fields] OR “severe acute respiratory syndrome coronavirus 2”[All Fields]) AND vaccine terms (“vaccin”[All Fields] OR “vaccine”[All Fields] OR “vaccines”[All Fields] OR “vaccination”[All Fields] OR “vaccinable”[All Fields] OR “vaccinal”[All Fields] OR “vaccinate”[All Fields] OR “vaccinated”[All Fields] OR “vaccinates”[All Fields] OR “vaccinating”[All Fields] OR “vaccinations”[All Fields] OR “vaccination’s”[All Fields] OR “vaccinator”[All Fields] OR “vaccinators”[All Fields] OR “immunization”[All Fields] OR “immunizations”[All Fields] OR immuniz* OR immunis*) AND survey terms (“survey”[All Fields] OR “surveys”[All Fields] OR “survey’s”[All Fields] OR “surveyed”[All Fields] OR “surveying”[All Fields] OR “questionnaire”[All Fields] OR “questionnaires”[All Fields]) OR “poll”[All Fields]).

All research articles published in the English language between December 1, 2019, and June 1, 2023, were collected using the search terms mentioned above.

### 2.2. Selection and Eligibility Criteria

Collected research articles were imported into COVIDENCE, a screening and data extraction tool for conducting systematic reviews [16]. COVIDENCE automatically removed all duplicate articles, yielding a set of studies to review. Subsequently, abstract and full-text screening were conducted by HM and SIT independently conducted on the resultant pool of articles and conflicts were addressed through unanimous consensus. Articles were included in this systematic review and meta-analysis based on the following inclusion criteria 1) they investigated COVID-19 vaccine acceptance, willingness, and uptake, 2) they were original cross-sectional survey studies, and 3) if they were conducted in China. The exclusion criteria were studies assessing 1) wrong outcome of interest, 2) willingness-to-pay or conditional vaccine acceptance, 3) non-COVID-19 vaccine acceptance, 4) continuous variables as they adopted different ranges of responses, and 5) studies conducted outside of China. Editorials, intervention studies, reviews, commentaries, letters, and qualitative research articles were also excluded for the purpose of this review.

### 2.3. Data Extraction

Data was extracted from all studies that met the inclusion criteria. The following data was collected onto an Excel sheet: article title, author name, date of publication, country, study design, sample size, sampling method, response rate, age of participants, data collection period, study objective, and type of publication (journal or pre-print service). To meet the goals of this systematic review, the following outcomes were also extracted: 1) overall COVID-19 vaccine acceptance/willingness (total sample), 2) overall COVID-19 vaccine uptake (total sample), 3) COVID-19 vaccination uptake across different subgroups (age, gender, income, and education), 4) COVID-19 vaccination acceptance/willingness, uptake, unsure, across different population groups (adults, healthcare workers, patients with chronic diseases, pregnant women, university students, and parents), and 5) reasons for COVID-19 vaccination refusal.

### 2.4. Risk of Bias Assessment

The NHLBI Quality Assessment Tool for Observational Cohort and Cross-Sectional Studies was used to assess the methodological risk of bias for all studies included in this systematic review and meta-analysis [17]. 14 criteria were used within this tool to answer the following questions for each study: (1) Was the research question or objective in this paper clearly stated? (2) Was the study population clearly specified and defined? (3) Was the participation rate of eligible persons at least 50%? (4) Were all the subjects selected or recruited from the same or similar populations (including the same time period)? Were inclusion and exclusion criteria for being in the study prespecified and applied uniformly to all participants? (5) Was a sample size justification, power description, or variance and effect estimates provided? (6) For the analyses in this paper, were the exposure(s) of interest measured prior to the outcome(s) being measured? (7) Was the timeframe sufficient so that one could reasonably expect to see an association between exposure and outcome if it existed? (8) For exposures that can vary in amount or level, did the study examine different levels of the exposure as related to the outcome (e.g., categories of exposure or exposure measured as a continuous variable)? (9) Were the exposure measures (independent variables) clearly defined, valid, reliable, and implemented consistently across all study participants? (10) Was the exposure(s) assessed more than once over time? (11) Were the outcome measures (dependent variables) clearly defined, valid, reliable, and implemented consistently across all study participants? (12) Were the outcome assessors blinded to the exposure status of participants? (13) Was the loss to follow-up after baseline 20% or less? (14) Were key potential confounding variables measured and adjusted statistically for their impact on the relationship between exposure(s) and outcome(s)?

For each question, there were three answer options: 1) Criteria satisfied, 2) criteria unsatisfied, and 3) Not applicable. Overall study quality ratings were assigned based on author rating, including three levels: “good,” “fair,” and “poor.” Each study was initially assigned a “good” rating and was demoted by one level per unsatisfied criteria.

### 2.5. Statistical analysis

Data organization and analysis from the included peer-reviewed studies were conducted using Microsoft Excel and Review Manager 5.4.1, respectively. All figures were created using Review Manager 5.4.1. Microsoft Excel was used to compute data in the tables and the pooled uptake, acceptance, and unsure rates.

#### 2.5.1. COVID-19 vaccine uptake outcome measures

The primary outcome was the uptake rates of COVID-19 vaccination amongst the population surveyed in China. The uptake status of Chinese residents was categorized into two groups: 1) Yes and 2) No. The pooled uptake rates were categorized for the overall study participants and divided according to respective population groups, such as 1) adults, 2) healthcare workers, 3) patients with chronic diseases, 4) pregnant women, 5) university/college students, and 6) parents. The category labelled “other” populations refers to individuals that cannot be categorized into the six categories mentioned above. These types of populations included caregivers, cold-chain workers, and the general public.

#### 2.5.2. COVID-19 vaccine acceptance outcome measures

Another outcome of interest was the rate of acceptance of COVID-19 vaccination. Acceptance was defined as study participants willing to/accepting/intending to receive the COVID-19 vaccination if the opportunity was made available. Acceptance was stratified based on the survey responses reported in the selected studies: 1) Yes/definitely; 2) Unsure/Do not know; 3) No/Definitely not. The pooled acceptance rates were reported for the overall study participants and for the different population groups outlined above.

#### 2.5.3. Data synthesis

Data were synthesized in the form of forest plots to compare the likelihood of uptake of the COVID-19 vaccination service across four population sub-groups. The sub-groups were created based on the following socio-demographic characteristics: (1) Age; (2) Gender; (3) Income; and (4) Education. Age was divided into five categories: (1) 18-29; (2) 30-39; (3) 40-49; (4) 50-59; (5) ≥60. Gender was reported as two categories: (1) Male; (2) Female. Income also had two categories: (1) <5000; (2) ≥ 5000. The income was reported as the local currency of China, Renminbi (RMB). Education had two categories: (1) High School or below; (2) Bachelor’s/college or above.

Each socio-demographic group had a reference range, and the Odds Ratio (OR) and 95% confidence interval (CI) were reported for each range. An OR >1 indicates a higher likelihood to uptake compared to the reference group. The pooled uptake outcome was reported if the following criteria were met: 1) two or more studies reported the same COVID-19 vaccine uptake outcome measure, and 2) the I^2^ statistic ≤60%.

An I^2^ value of >60% was considered to be heterogeneous on a statistical level. To meta-analyse the data, an inverse variance statistical analysis was conducted with a random-effects model Meta-analysis was only conducted if there were two or more studies in the pool. Additionally, a P-value of <0.05 was considered to be statistically significant. A sensitivity analysis was conducted to examine outliers for the OR and 95% CI values for COVID-19 vaccination acceptance.

### 2.6. Quality of evidence evaluation

The quality of evidence gathered in the meta-analysis was assessed using Cochrane’s GRADE approach [18]. The quality rating had four levels: “high,” “moderate,” “low,” and “very low.” All of the resulting outcomes started with a “high” rating and were demoted one level per unsatisfied criteria

#### 2.6.1. Publication bias

Publication bias was assessed through the analysis of funnel plot symmetricity. Publication bias was not assessed for quantitative analyses with <5 studies due to a lack of statistical power. The funnel plot was created for two forest plots (Age and Gender) through the Review Manager 5.4.1 statistical analysis software. To test for the presence of publication bias, the funnel plots were visually analyzed and interpreted.

## 3. Results

### 3.1. Identification and Selection of Studies

A total of 2420 studies were found through the literature searches 881 were found through the Web of Science journal, 781 through Embase, 593 through Scopus, 144 through EBSCO, and 21 through PubMed. 1161 duplicates were found and excluded. A total of 1259 studies were available for screening. 1075 studies were excluded in the abstract screening, and 184 studies remained for full-text screening. A total of 137 studies were excluded after the full-text screening: 3 for having the wrong study setting, 115 for analyzing the wrong outcome measure, 3 for using the wrong comparator, 2 for having the wrong intervention, 10 for having the wrong study design, and 4 for studying the wrong population. Finally, 47 studies were available for data extraction. A flowchart of study selection according to PRISMA guidelines can be seen in Figure 1 below.

**Figure 1.**
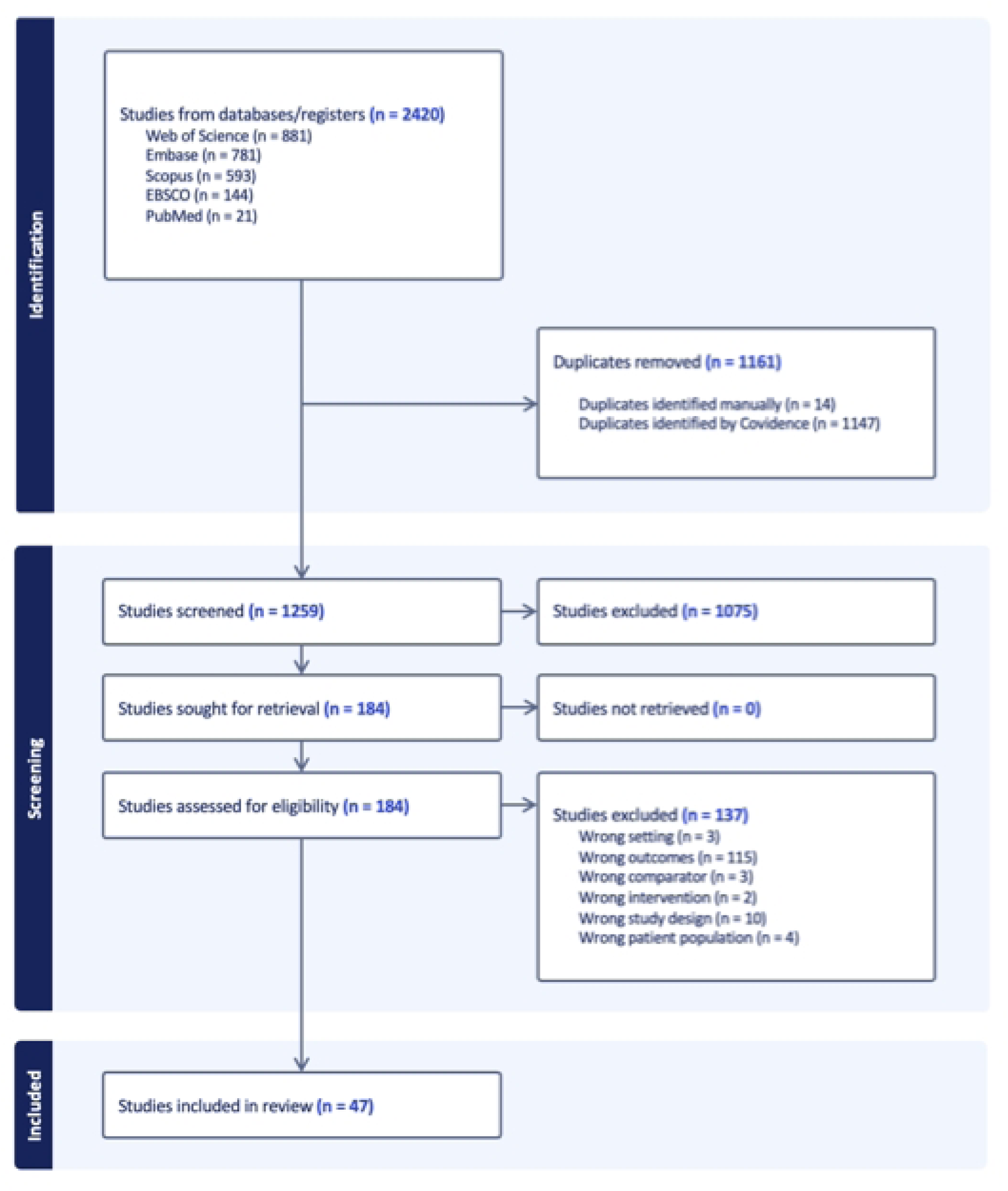
PRISMA Flowchart of Study Selection

### 3.2. Study Characteristics

The main characteristics of the studies included in the systematic review and meta-analysis are presented in Table 1, and data collection times for the selected studies ranged from March 2020 to December 2022. The age ranges of the study participants ranged from 12-75 years. All of the included studies were conducted in China, and the sample sizes ranged from 208 to 96498 participants. While some studies did not report the response rates of the surveys used to obtain data, the lowest response rate recorded was 55%, and the highest was 99.3%. There were 19 studies that did not report the sampling method used to select study participants. 13 studies utilized convenience sampling, five studies used snowball sampling, two used multi-stage sampling, five used random sampling, two used cluster sampling, and 1 study used purposive sampling. All 47 articles were cross-sectional studies published in peer-reviewed journals, and there were no studies published in a pre-print service.

**Table 1.** Overview of studies included in systematic review and meta-analysis.

| # | Author/<br>publication year | Study Design | Country | Data collection time | Age range<br>(years) | Sample size (n) | Response<br>rate<br>(%) | Sampling<br>method | Publication |
| --- | --- | --- | --- | --- | --- | --- | --- | --- | --- |
| 1 | Chang et al. 2023<br>[19] | cross-sectional | China | December 2021 to April 2022 | ≥18 | 261 | NR | NR | Journal |
| 2 | Chan et al. 2021<br>[20] | cross-sectional | China | 31 January 2021 and 15 February 2021 | ≥18 | 660 | 75 | Convenience | Journal |
| 3 | Chen et al. 2021<br>[21] | cross-sectional | China | May to June 2020 | ≥18 | 3195 | NR | Snowball | Journal |
| 4 | Feng et al. 2021 [22] | cross-sectional | China | November 17, 2020, to January 28, 2021 | ≥20 | 3703 | NR | Multi-stage | Journal |
| 5 | Fu et al. 2022 [23] | cross-sectional | China | August to November 2021 | ≥18 | 343 | NR | NR | Journal |
| 6 | Gan et al. 2021 [24] | cross-sectional | China | 23 October to 10 November 2020 | 18 to 74 | 1009 | NR | Convenience | Journal |
| 7 | Hao et al. 2022 [25] | cross-sectional | China | April 2021 to June 2021 | 18 to 60 | 621 | 93.67 | Convenience | Journal |
| 8 | Hou et al. 2022 [26] | cross-sectional | China | September 14 to November 18, 2021 | ≥18 | Caregivers: 2588<br>HCW: 1700 | 84.8 | Multi-stage | Journal |
| 9 | Huang et al. 2021 [27] | cross-sectional | China | January and February 2021 | 18-65 | 2740 | NR | NR | Journal |
| 10 | Huang et al. 2023 [28] | cross-sectional | China | May 1 to 31, 2022 | ≥18 | 11565 | 77.81 | NR | Journal |
| 11 | Huang et al. 2022 [29] | cross-sectional | China | First survey: January 2021<br>Second survey: June of 2021 | NR | NR | 97 | NR | Journal |
| 12 | Hu et al. 2022 [30] | cross-sectional | China | 1 and 20 May 2021 | ≥18 | 359 | NR | NR | Journal |
| 13 | Kong et al. 2022 [31] | cross-sectional | China | 19 June 2021 to 23 June 2021 | ≥18 | 786 | NR | Snowball | Journal |
| 14 | Li et al. 2022 [32] | cross-sectional | China | May 19 to June 18, 2021 | ≥18 | 2176 | NR | NR | Journal |
| 15 | Li et al. 2022 [33] | cross-sectional | China | 20 to 27 January 2022 | ≥18 | 11141 | NR | Convenience | Journal |
| 16 | Li et al. 2021 [34] | cross-sectional | China | June 2020 | ≥18 | 2377 | 88.04 | Snowball | Journal |
| 17 | Lin et al. 2020 [35] | cross-sectional | China | 1–19 May 2020 | ≥18 | 3541 | NR | NR | Journal |
| 18 | Li et al. 2022 [36] | cross-sectional | China | 10 and 28 June 2021 | 18-23 | 721 | 83.9 | Random | Journal |
| 19 | Li et al. 2022 [37] | cross-sectional | China | 16 August to 28 October 2021 | 12-17 | 2048 | NR | Cluster | Journal |
| 20 | Liu et al. 2022 [38] | cross-sectional | China | 16 December 2020 and 21 December 2020 | ≥18 | 244 | 96 | Random | Journal |
| 21 | Liu et al. 2023 [39] | cross-sectional | China | NR | ≥18 | 96498 | 80 | Random | Journal |
| 22 | Li et al. 2021 [40] | cross-sectional | China | January 20 to February 20, 2021 | ≥18 | 1779 | 95.5 | Convenience | Journal |
| 23 | Luk et al. 2021 [41] | cross-sectional | China | 9 to 23 April 2020 | ≥18 | 1501 | 61.3 | NR | Journal |
| 24 | Lv et al. 2023 [42] | cross-sectional | China | January 2022 to March 2022 | ≥18 | 1424 | 91.6 | Convenience | Journal |
| 25 | Mo et al. 2022 [43] | cross-sectional | China | October and November 2020 | ≥18 | 1733 | NR | NR | Journal |
| 26 | Pan et al. 2022 [44] | cross-sectional | China | 26 April to 10 May 2022 | ≥18 | 449 | 97.3 | Convenience | Journal |
| 27 | Qin et al. 2023 [45] | cross-sectional | China | 26 December to 31 December 2022 | ≥18 | 42565 | 99 | Cluster | Journal |
| 28 | Qin et al. 2021 [46] | cross-sectional | China | November 12, 2021 | ≥18 | 1724 | NR | NR | Journal |
| 29 | Song et al. 2022 [47] | cross-sectional | China | January 29 to February 4, 2021. | ≥18 | 2244 | NR | Snowball | Journal |
| 30 | Sun et al. 2022 [48] | cross-sectional | China | December 1 to December 9, 2020 | ≥18 | 3047 | NR | NR | Journal |
| 31 | Tao et al. 2021 [49] | cross-sectional | China | November 13 to 27, 2020. | ≥18 | 1392 | NR | NR | Journal |
| 32 | Wang et al. 2021 [50] | cross-sectional | China | January 10 to January 22, 2021 | 18-60 | 8742 | 91.7 | NR | Journal |
| 33 | Wang et al. 2021 [51] | cross-sectional | China | 10 January to 22 January 2021 | 18-60 | 2386 | NR | Convenience | Journal |
| 34 | Wang et al. 2020 [52] | cross-sectional | China | March 2020 | ≥18 | 2,058 | 98 | Random | Journal |
| 35 | Wang et al. 2021 [53] | cross-sectional | China | November 2020 to January 2021 | $\geq 18$ | 7259 | NR | Convenience | Journal |
| 36 | Wang et al. 2021 [54] | cross-sectional | China | January 2021 | $\geq 18$ | 2580 | NR | NR | Journal |
| 37 | Wong et al. 2021 [55] | cross-sectional | China | July to August, 2020 | $\geq 18$ | 1200 | 55 | Random | Journal |
| 38 | Wu et al. 2022 [56] | cross-sectional | China | July 10, 2021 | $\geq 18$ | 29925 | NR | Snowball | Journal |
| 39 | Wu et al. 2022 [57] | cross-sectional | China | May to June 2021 | 60-75 | 1,067 | 97 | Convenience | Journal |
| 40 | Wu et al. 2022 [58] | cross-sectional | China | 4 April to 18 April 2021 | $\geq 18$ | 1126 | NR | NR | Journal |
| 41 | Xie et al. 2023 [59] | cross-sectional | China | NR | $\geq 60$ | 951 | NR | NR | Journal |
| 42 | Yin et al. 2021 [60] | cross-sectional | China | March 25, 2021 to April 2, 2021 | $\geq 18$ | 23940 | NR | Convenience | Journal |
| 43 | Zhang et al. 2021 [61] | cross-sectional | China | August 16–20, 2021 | $\geq 22$ | 631 | NR | NR | Journal |
| 44 | Zhang et al. 2022 [62] | cross-sectional | China | 26 and 31 October 2021 | $\geq 18$ | 2329 | 85.8 | Purposive | Journal |
| 45 | Zhang et al. 2023 [63] | cross-sectional | China | Round1: 1 to 7 September 2020<br>Round 2: 26 to 31 October 2021 | $\geq 18$ | Round 1: 208<br>Round 2: 229 | 85.8 | NR | Journal |
| 46 | Zhao et al. 2021 [64] | cross-sectional | China | 29 January 2021 to 26 April 2021 | $\geq 18$ | 34041 | 99.3 | Convenience | Journal |
| 47 | Zhou et al. 2021 [65] | cross-sectional | China | 1 March 2021 and 10 April 2021 | 18-65 | 3940 | NR | Convenience | Journal |

HCW = Healthcare workers; NR = Not reported

#### 3.2.1. Participant characteristics

For qualitative data synthesis in this systematic review, data were collected from a total of 327,046 participants. For the meta-analysis, data from 130,441 participants was used.

#### 3.2.2. COVID-19 vaccine uptake outcome measures

Amongst the 47 studies included in the review, ten studies (21.3%) reported the OR and 95% CI for the uptake of COVID-19 vaccines for different socio-demographic factors. The remaining 37 studies reported the OR and 95% CI for the acceptance/willingness to receive COVID-19 vaccines. For the purpose of meta-analysis, data was only used for uptake of vaccination because it was deemed to be a more robust metric for assessing vaccination habits compared to acceptance/willingness which may be translatable Thus, ten studies were included in the meta-analysis.

### 3.3. Risk of bias (quality) analysis for included studies

Results from the risk of bias assessment are reported in S1 Table. After conducting the quality assessment, the majority of the studies were rated as good quality (42/47), while the remaining studies were rated as fair quality (5/47). These five studies were demoted to having fair quality as they failed to report the outcome measures (dependent variables) consistently and clearly. Within these studies, the socio-demographic groups were not completely reported or the ranges differed substantially from the remaining study pool [31, 45, 60, 61, 65).

### 3.4. Estimated percentage of acceptance, unsure, and uptake outcomes

Based on data collected from all 47 studies, the acceptance, unsure, and uptake rates of COVID-19 vaccination were reported in Table 2. 35 studies investigated the rate of acceptance/willingness to receive COVID-19 vaccines, 8 studies reported the number of participants unsure about receiving the vaccine, and 21 studies reported the uptake rates of COVID-19 vaccines. The overall acceptance rate of COVID-19 vaccination in China was calculated to be 59.5%. In comparison, 20.9% of the sampled population was found to be unsure about COVID-19 vaccines. The estimated COVID-19 vaccination uptake rate was found to be 69.9% amongst the sampled population. The acceptance, unsure, and uptake rates varied across each population group. The highest vaccination acceptance rate was found to be for university/college students (87.4%), while the lowest rate was observed for patients with chronic diseases (42.6%). The highest unsure rate was reported for patients with chronic diseases (48.3), while university/college students had the lowest (2.5%). Additionally, the highest COVID-19 vaccination uptake rate was observed for Chinese adults (92%), and the lowest was seen for patients with chronic diseases (43.5%).

**Table 2.** Summary of estimated acceptance, unsure, uptake rates.

| Population groups | Acceptance of COVID-19 Vaccines |  |  |  | Unsure |  |  |  | Uptake of COVID-19 Vaccines |  |  |  |
| --- | --- | --- | --- | --- | --- | --- | --- | --- | --- | --- | --- | --- |
|  | No. of studies | No. of participants | Total no. of accepting | Estimated acceptance (%) | No. of studies | No. of participants | Total no. of unsure | Estimated unsure (%) | No. of studies | No. of participants | Total no. of uptake | Estimated uptake (%) |
| Overall | 35 | 135657 | 80,705 | 59.5 | 8 | 35332 | 7400 | 20.9 | 21 | 258124 | 180358 | 69.9 |
| Adults | 7 | 39043 | 28,360 | 72.6 | 4 | 29211 | 5101 | 17.5 | 3 | 44302 | 40774 | 92.0 |
| Healthcare Workers | 5 | 12806 | 9796 | 76.5 | 0 | NA | NA | NA | 3 | 8453 | 6183 | 73.2 |
| Patients with underlying health conditions | 3 | 4526 | 1,926 | 42.6 | 2 | 3400 | 1642 | 48.3 | 4 | 4,768 | 2076 | 43.5 |
| Pregnant women | 1 | 1,392 | 1077 | 77.4 | 0 | NA | NA | NA | 0 | NA | NA | NA |
| University/college students | 1 | 721 | 630 | 87.4 | 1 | 721 | 18 | 2.5 | 2 | 990 | 762 | 76.9 |
| Parents | 4 | 10152 | 8,863 | 87.3 | 0 | NA | NA | NA | 3 | 26994 | 23184 | 85.9 |
| Others | 14 | 67017 | 30,053 | 44.8 | 1 | 2000 | 639 | 31.9 | 6 | 172617 | 107379 | 62.2 |
| NA = Not applicable |  |  |  |  |  |  |  |  |  |  |  |  |

### 3.5. Reasons for COVID-19 vaccine refusal

The most commonly cited reasons for refusing the COVID-19 vaccine among the study samples are presented in Table 3. The reasons are shown for studies included in the meta-analysis. Amongst these ten studies, 7 (70%) investigated the factors that affect the Chinese population’s decision to uptake COVID-19 vaccination services and the reasons for refusing vaccines. Among these 7 studies that report this data, 6 (85.7%) utilized descriptive statistics to showcase the percentage of total participants that refused the COVID-19 vaccine due to the stated reasons.

**Table 3.** Reasons for vaccine refusal in studies included in meta-analysis.

| Reference | Common reasons for refusal | No. of refusers (% of sample pool) |
| --- | --- | --- |
| Fu et al. 2022 [23] | Worried about vaccine safety | 67% |
| Hou et al. 2022 [26] | Lack of evidence about vaccine safety | 82.26 |
| Huang et al. 2021 [27] | NR | NR |
| Huang et al. 2023 [28] | Fear of side-effects | 53.78 |
| Li et al. 2021 [34] | vaccine's safety | 37.6% |
| Qin et al. 2023 [45] | NR | NR |
| Song et al. 2022 [47] | Concerns about safety and efficacy | NR |
| Wu et al. 2022 [56] | Skepticism about vaccine | 47.2 |
| Xie et al. 2023 [59] | NR | NR |
| Zhao et al. 2021 [64] | Concerns about vaccine safety | 71.9 |
| NR = Not reported |  |  |

The majority of the studies found that concern about COVID-19 vaccine safety was the most common area of concern amongst the sampled population. Other most commonly stated reasons for vaccine refusal were fear about its side effects and the overall efficacy of the vaccine against COVID-19. Hou et al. (2022) found that out of the proportion of participants that refused to receive the COVID-19 vaccine, 82.26% refused to get immunized due to a lack of information regarding vaccine safety.

### 3.6. Sensitivity analysis

Two sensitivity analyses were conducted. The first sensitivity analysis was the exclusion of uptake outcome values (OR, 95% CI), whereby the socio-demographic groups (age and income) were substantially far apart from the reference group. Uptake outcome values were also excluded for studies that had incongruent reference groups. This sensitivity was conducted in 4 out of 14 studies that reported OR and 95% CI for socio-demographic groups and were excluded from the meta-analysis: [19, 31, 42, 61].

The second sensitivity analysis was the exclusion of Fu et al. [23] from the meta-analysis for income. This was done in order to assess whether this study was a detrimental outlier and played a significant role in the overall uptake outcome.

### 3.7. Quality of evidence

GRADE assessment was conducted for all four meta-analyses (Age, gender, income, education) and is presented in S2 - S5 Tables. One meta-analysis for OR - Income was assessed to have high quality. Two meta-analyses for OR - Age and OR - Gender were assessed to have moderate quality. One meta-analysis for education was analyzed to be low in quality according to the GRADE criteria.

### 3.8. Meta-analysis

Forest plots were created for four socio-graphic groups: (1) Age; (2) Gender; (3) Income; (4) Education.

#### 3.8.1. Odds Ratio – Age

Five studies with fifteen outcomes displayed varied results for OR of vaccine uptake. Statistical pooling was inappropriate for the age group because of statistical heterogeneity (I^2^ = 77%) (Figure 2).

**Figure 2.**
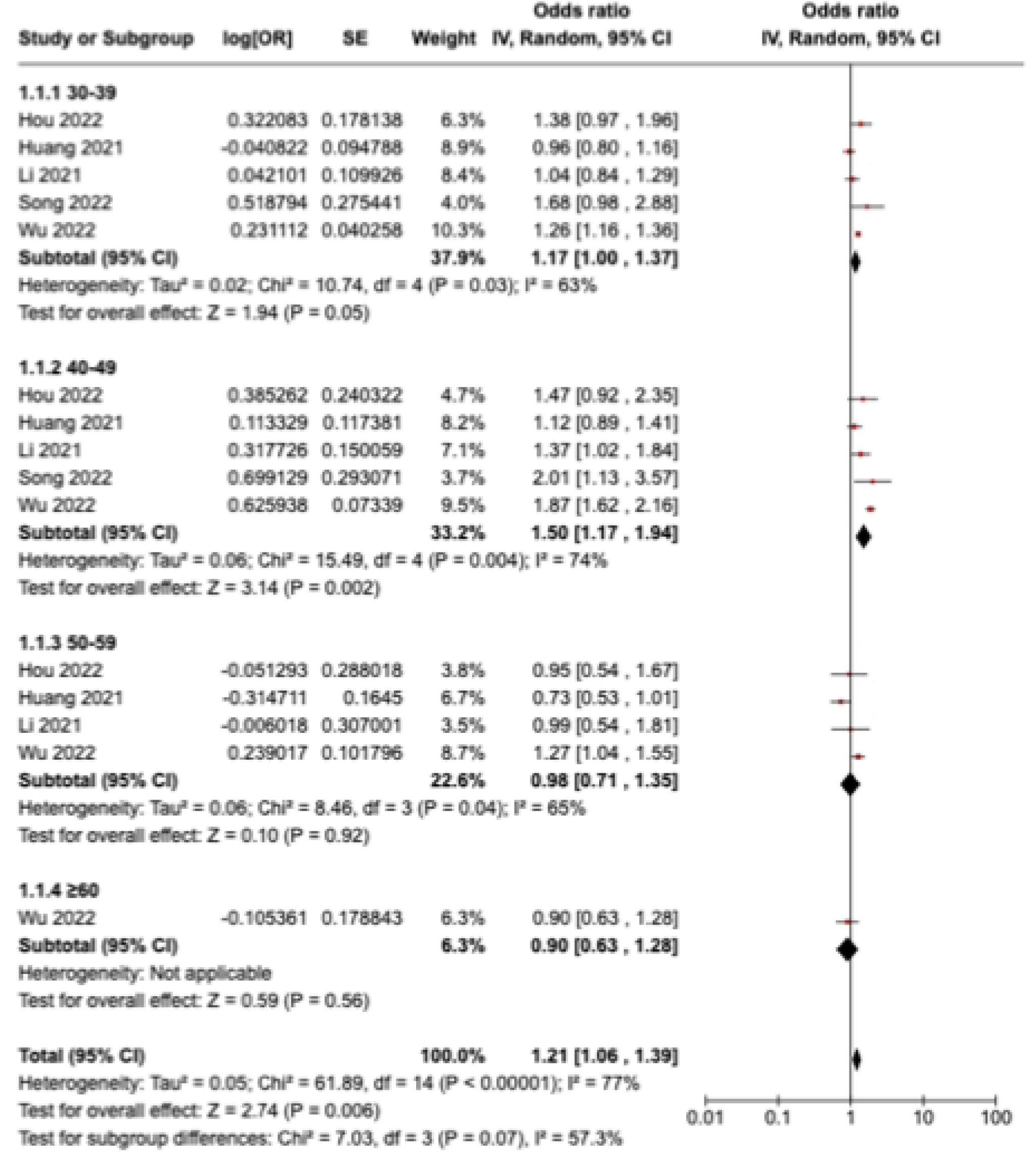
Age Odds Ratio for sub-groups uptaking COVID-19 vaccine compared to reference group

#### 3.8.2 Odds Ratio – Gender

Nine studies with nine outcomes displayed varied results for OR of vaccine uptake. Statistical pooling was inappropriate for the gender group because of statistical heterogeneity (I^2^ = 97%) (Figure 3). The OR value and 95% CI for the uptake likelihood of male gender were statistically significant as the P-value was equal to 0.0001 and the data was homogeneous (I^2^ = 0%). Males were more likely to uptake COVID-19 vaccination (OR=1.17, 95% CI:1.08-1.27) (Figure 3).

**Figure 3.**
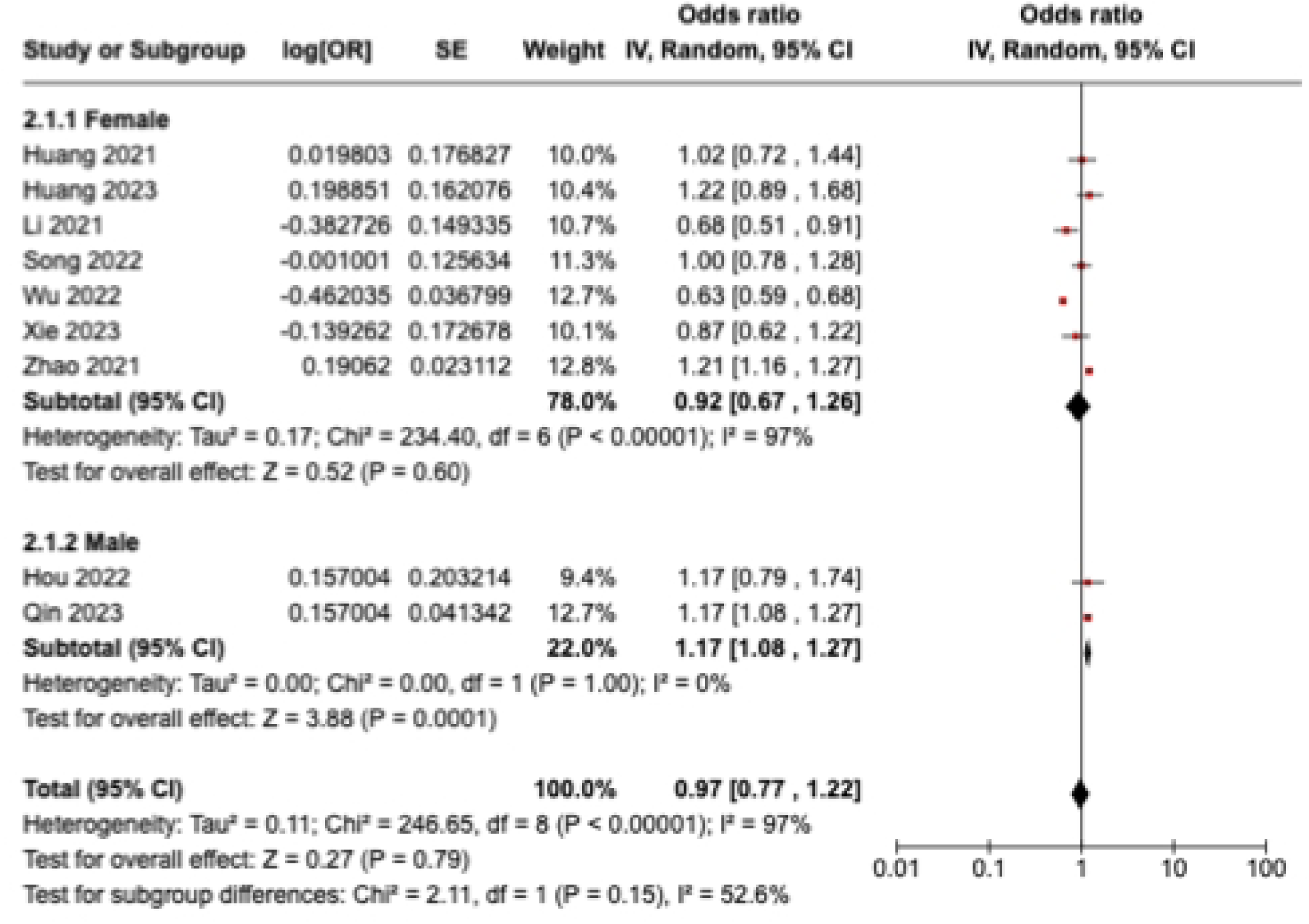
Gender Odds Ratio for sub-groups uptaking COVID-19 vaccine compared to reference group

#### 3.8.3 Odds Ratio – Income

Three studies with three outcomes had different results for OR of vaccine uptake. Li et al. [40], Xie et al. [59], and Zhao et al. [64] reported an increase in the likelihood of uptaking COVID-19 vaccination for individuals with a monthly income or more than or equal to 5000 RMB. The reference group for this analysis included participants with a monthly income of less than 5000 RMB. Statistical pooling was appropriate due to statistical homogeneity (I^2^ = 0%) (Figure 4). There was an overall increase in the likelihood of vaccine uptake (OR=1.08, 95% CI:1.02-1.14), and it was statistically significant (P-value = 0.005).

**Figure 4.**
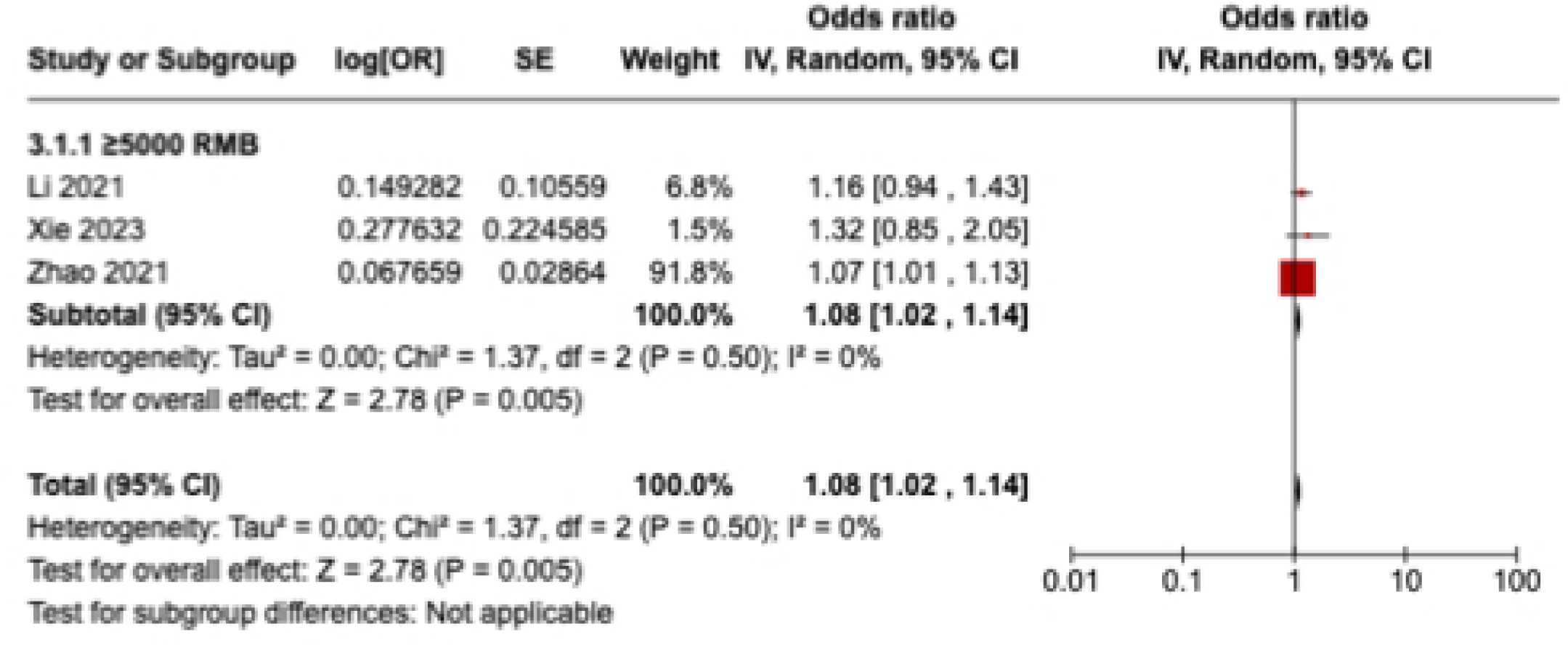
Income Odds Ratio for sub-groups uptaking COVID-19 vaccine compared to reference group

#### 3.8.4 Odds Ratio – Education

Four studies with four outcomes displayed varied results for OR of vaccine uptake. Statistical pooling was inappropriate for the gender OR values because of statistical heterogeneity (I^2^ = 88%) (Figure 5).

**Figure 5.**
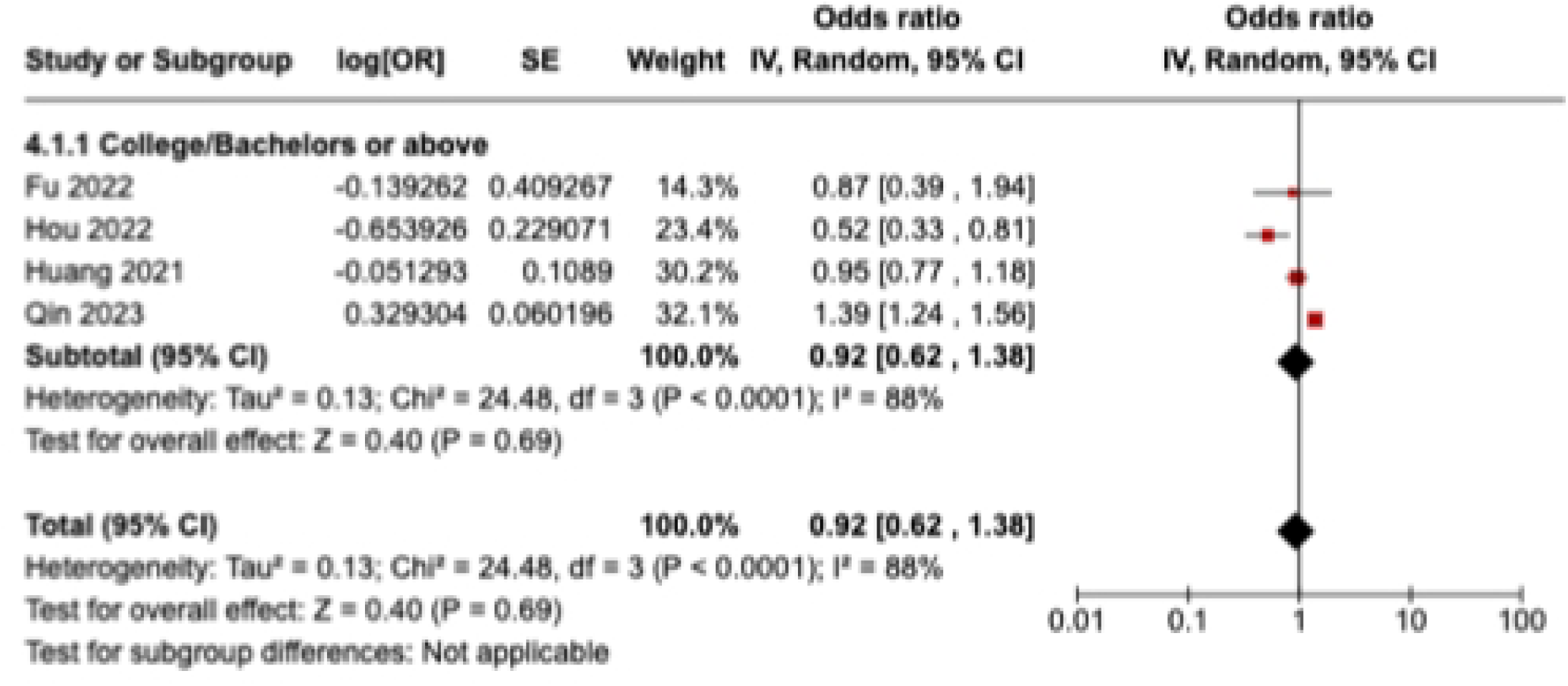
Education Odds Ratio for sub-groups uptaking COVID-19 vaccine compared to reference group

### 3.9. Publication bias

The funnel plots generated to detect publication bias are presented in S1 - S2 Fig. As the quantitative analysis for income and education socio-demographic groups were derived from 3 and 4 studies, respectively, funnel plots were not assessed for publication bias due to a lack of statistical power.

#### 3.9.1. Odds Ratio – Age

The funnel plot for the age socio-demographic group is presented in S1 Fig. The majority of the studies are plotted near the average (dotted line), which indicates high precision.

Therefore, no publication bias was detected visually.

#### 3.9.2. Odds Ratio – Gender

The funnel plot for the gender socio-demographic group is presented in S2 Fig. The majority of the studies are plotted near the average (dotted line), which indicates high precision. Therefore, no publication bias was detected visually.

#### 3.9.3. Odds Ratio – Income

A funnel plot was not created for the Income funnel plot as there were insufficient studies (<5), lowering the statistical power of publication bias analysis.

#### 3.9.4. Odds Ratio – Education

A funnel plot was not created for the Education funnel plot as there were insufficient studies (<5), lowering the statistical power of publication bias analysis.

## 4. Discussion

The purpose of this systematic review and meta-analysis was to extrapolate the acceptance and uptake of COVID-19 vaccination among Chinese residents. This paper also reports common reasons for vaccine refusal among the Chinese population, along with factors influencing the decision to vaccinate. Currently, there are no systematic reviews that identify the uptake and willingness to vaccinate oneself across different population groups in China. This paper reports for the first time vaccination behaviours across different groups (adults, healthcare workers, patients with chronic diseases, pregnant women, university students, and parents) and meta-analyzes the likelihood of specific socio-demographic factors (age, gender, income, and education) influencing uptake COVID-19 vaccines.

### 4.1. Acceptance of COVID-19 vaccines

The analysis conducted in this review revealed that more than half of the overall Chinese population (59.5%) was accepting of the COVID-19 vaccination and believed that it was necessary to control the spread of the COVID-19 pandemic. However, this review also indicated that there is variation regarding the rate of acceptance of COVID-19 vaccines across different population groups. The highest rate of acceptance in China was observed amongst university/college students (87.4%). Greater exposure to the scientific curriculum as per the academic curriculum could potentially explain the higher acceptance rate among students, as they might be more aware of the efficacy of vaccines.

The review also found that the acceptance of COVID-19 vaccines was lower amongst healthcare workers (HCWs) compared to other groups, such as students and pregnant women. These HCWs were not willing to receive COVID-19 vaccines despite having access is crucial to increase the rate of acceptance and willingness of COVID-19 vaccination among the healthcare workforce as they provide care to patients and are a high-risk group for spreading infectious diseases [40].

#### 4.1.1. Vaccine hesitancy among those with underlying health conditions

Furthermore, this systematic review found that the lowest acceptance rate of COVID-19 vaccination was amongst patients with chronic diseases (42.6%). These patients were diagnosed with diseases such as asthma, HIV/AIDS, cancer, and hypertension [39, 27, 23, 19, 20]. Comparing these acceptance rates with those of healthy adults (72.6%), this is a substantial difference. There is also a difference in uptake rates, with 92% of healthy adults receiving COVID-19 vaccines compared to 43.5% of patients with underlying disease. These observable differences could be due to concern about the side effects of COVID-19 vaccines on underlying conditions, many of which are rooted in misinformation (citation). It is important to increase knowledge dissemination regarding the safety of COVID-19 vaccines and potential side effects, to fight disinformation and gain patient confidence.

Another study by Lv et al. [42]found that people with underlying conditions were more likely to be of old age. Lv et al. [42] found that older people with underlying conditions had difficulty grasping, assessing and analyzing media content, which further added to the development of misconceptions. People with health conditions also had a higher likelihood of dealing with anxiety and depression [42]. Chinese residents with moderate or high levels of anxiety had a significantly higher vaccine refusal rate [42]. Moreover, the anxiety suppressors being taken by these patients were found to have a correlation with increasing vaccine hesitancy amongst those with underlying conditions [42]. These findings point to the intersectionality of multifarious factors that lead to vaccine hesitancy and consequent refusal. It is vital to understand these factors at length to make informed decisions about ways to increase the overall COVID-19 vaccine acceptable rate in China.

### 4.2. Uncertainty about COVID-19 vaccines

This review uncovered that a substantial amount of the Chinese population is unsure about receiving the COVID-19 vaccine. Overall, the rate of unsure participants was 20.9%. However, there is a lot of variation amongst different population groups. The highest percentage of people unsure about COVID-19 vaccination were found to be patients with chronic diseases (48.3%). In fact, 17.5% of the adult population in China was unsure regarding the safety and efficacy of COVID-19 vaccines. On the other hand, the lowest rate of uncertainty was found within students enrolled in colleges or universities (2.5%). This goes to show that education is an important determinant of vaccine hesitancy.

No articles included in this review reported quantitative data for HCWs and pregnant women, which indicates a knowledge gap regarding vaccination perceptions and behaviours within these groups.

### 4.3. Uptake of COVID-19 vaccines

The overall prevalence of COVID-19 vaccine uptake among the Chinese population was 69.9%. The findings revealed notable differences in the rates of uptake of COVID-19 vaccines across population groups. The highest uptake rate was found to be for Chinese adults (92%). Some reasons for this exceptionally high rate could be due to the serious impacts of the COVID- 19 pandemic on their daily lives. Notably the disruptions regarding work, travel restrictions, and severe economic losses [21]. Consequently, this might have led to dynamic changes where adults were more informed about preventative measures such as COVID-19 vaccines, increasing their rate of uptake. However, the rate of uptake of COVID-19 vaccination was lower for healthcare workers (73.2%). The most common reason for this low uptake rate is the aforementioned lack of confidence in the safety and efficacy of COVID-19 vaccines [50]. More importantly, some HCWs did not believe that the vaccine was a reliable method of tackling the spread of the COVID-19 pandemic [50]. The uptake rates were also vastly different for the type of HCWs. Administration employees in healthcare environments were more likely to be skeptical about the vaccines, whereas doctors and nurses had a higher uptake rate [50]This could be due to the nature of the jobs. HCWs, such as doctors, have a stronger medical background and are more knowledgeable about the benefits of vaccines. In comparison, the lowest uptake rate was observed for patients with chronic diseases in China. This is in accordance with the high level of concerns about vaccines and low acceptance observed earlier.

Males were also more likely to uptake COVID-19 vaccination services (OR=1.17; 95% CI:1.08 - 1.27) when compared to females in China. These results imply that males are 17% more likely to get vaccinated against COVID-19 compared to females. Given that P-value=0.0001, the findings were also statistically significant. Similar to acceptance/uncertainty regarding COVID-19 vaccines, reasons for the higher uptake rate amongst Chinese males could be due to a higher perception of the detrimental effects of COVID-19 and less observance of misconceptions

The quantitative analysis further highlighted important differences in COVID-19 vaccine uptake rates based on the income of Chinese residents. In terms of the local currency of China (RMB), it was observed that having a higher monthly income corresponded to a greater likelihood of uptake of COVID-19 vaccines. Specifically, Chinese residents with a monthly income ≥ 5000 RMB had higher uptake of vaccination (OR=1.08; 95% CI:1.02 - 1.14) when compared to those with a monthly income < 5000 RMB. This shows that having an income ≥ 5000 RMB results in an 8% higher likelihood of uptake of COVID-19 vaccines. As the P-value =0.005, the results were also statistically significant. Possible reasons for these findings include higher access to transportation, which can act as a facilitator to uptaking vaccination. Furthermore, a lower income could be associated with the geographic location or place of residence in China, whereby such individuals do not have access to an abundance of vaccination clinics. Individuals living in remote locations and lower-income households might not be able to receive the vaccine despite having the willingness. Also, income can function as a determinant of the daily schedules of Chinese residents. It is possible that having a lower household income increases stress and creates hectic daily schedules, which can lead to a lower likelihood of having the time to uptake COVID-19 vaccination.

#### 4.3.1. Varying degrees of vaccine hesitancy among different age groups

This analysis revealed a lower likelihood of COVID-19 vaccine uptake amongst older-aged individuals in China. Those within the 50-59 age bracket had a 2% lower uptake of COVID-19 vaccine. However, this percentage increases dramatically with age. Amongst those that were ≥60 years of age, there was a 10% lower uptake of vaccination. However, there were a low amount of studies reporting data for those over the age of 60, which is why the findings of this analysis need to be utilized with caution. There is a need to investigate in-depth recent changes in vaccination uptake rates amongst those over the age of 60 and specifically those that are 80 and above. The reason is that recent reports, as mentioned earlier, with the termination of restrictive policies from the zero-covid policy, show a steep rise in COVID-19 cases and mortality amongst those over the age of 80 in China.

A research study conducted by Smith et al. [66] has demonstrated clear findings showcasing a higher vaccination hesitancy/refusal and an extremely low coverage rate amongst Chinese elders. Among those aged 60 and above, the COVID-19 vaccination coverage rate declined with age. Only 48% of those that fall within the 70-79 year age bracket had received at least one dose of the COVID-19 vaccine [66]. Furthermore, amongst those that were ≥80 years, only 20% had received at least one vaccine [66]. The consequences of these low vaccination uptake rates are evident by looking at the case and fatality statistics. Amongst 5,906 deaths that were reported, 5,655 (96%) occurred among people in China aged 60 and above [66]. Moreover, of these deaths, 70% occurred among those that were unvaccinated against COVID-19 [66]. Notably, Smith et al. [66] found that within the 60 and above age group in China, the risk of death among unvaccinated individuals was 21.3 times higher than those that were vaccinated.

### 4.4. Strengths and Limitations

This study has several strengths and limitations. Firstly, this review was conducted according to the Cochrane guidance to ensure that the methodologies were robust and appropriate. Secondly, developing clear and focused inclusion/exclusion criteria allowed a clear scope for this systematic review. Thirdly, the database searches were conducted by clearly outlined and comprehensive search terms, which were applied to multiple databases (PubMed, EMBASE, Web of Science, EBSCO, and Scopus). Only peer-reviewed databases were screened, which allowed searching a broad spectrum of articles to gather detailed information on the research topic. Furthermore, the presence of publication biases and assessment of reporting biases were accounted for through the creation of funnel plots. These funnel plots were visually assessed for asymmetry to determine whether bias was present. No publication bias was detected for this systematic review and meta-analysis. This enhances the validity of this study and ensures transparency of the data reported.

However, there are some limitations to this study. Firstly, the research studies used to conduct this systematic review and meta-analysis derived data from cross-sectional analyses, which are referred to as snapshots of COVID-19 vaccination behaviours in each region. Cross-sectional studies can have diverse sampling methods such as convenience sampling, snowball sampling, multi-stage sampling, random sampling, cluster sampling, and purposive sampling. These differences can, to some extent, explain the differences observed in the acceptance, unsure, and uptake rates of COVID-19 vaccines across studies from a single country. Therefore, these results gathered need to be interpreted with caution as they might not be able to predict future changes in COVID-19 vaccination behaviours in China. Secondly, an essential limitation is with regard to the different methods used to acquire data regarding willingness and acceptance of COVID-19 vaccines amongst different population groups in China. Some studies utilized a binary response system within the questionnaires/surveys (yes/no), whereas other studies used a different breakdown of options (strongly agree/agree/neutral/disagree/strongly disagree) to assess attitudes regarding vaccination. Answers can differ based on the subjective perceptions of the participants surveyed within these studies. Therefore, these variables should be taken into consideration to ensure an accurate comparison of vaccination behaviours across the different studies included in this review.

### 4.5. Next steps of research

This review guides several next steps for prospective research studies. The current studies available for this review lacked an in-depth analysis of the perceptions of the general public regarding vaccines, as they only offered binary answers. Future research studies should conduct a more in-depth qualitative analysis by creating focus groups and conducting interviews with the sampled study participants. Employing such methods can help build a positive group dynamic and synergy where everyone is provided equal opportunity to share their opinions freely in a non-judgmental atmosphere. Not only is this method cost-effective, but it is efficient in gathering data to look beyond the numbers and truly understand the meaning behind the results. Future studies should also investigate the engagement of the Chinese population with media sources and other outlets to map out sources of misinformation and popular platforms in China and guide solutions to increase vaccination uptake rates. Also, future research studies regarding this topic should utilize a consistent model, such as during the design of the questionnaires, to enhance the precision and applicability of the findings.

### 4.6. Recommendations

There are several recommendations to bolster the overall acceptance and uptake of COVID-19 vaccination services in China. Firstly, a community health training model can be utilized as per the common reasons for refusal observed from this review. Lack of knowledge, negative attitudes towards vaccines, and misconceptions are possible reasons why the majority of the Chinese population is worried about the safety of COVID-19 vaccines. To address this, home visits and informational campaigns should be initiated that would aid in addressing the common misconceptions through one-on-one conversations between trained health professionals and the public. The public would be able to ask questions on the spot, which can lead to an overall increase in the acceptance of the campaign as well as vaccination services. A research study by Singh et al. [67] showcases that employing such strategies led to an overall increase in vaccination coverage rates from 21 to 33%.

Secondly, incentivizing the uptake of COVID-19 vaccines is an effective tool to improve vaccination coverage. In particular, this method can be helpful in targeting the rural and remote regions of China as they might be more willing to accept the notion due to the incentives set in place [67]. Monetary incentives will help overcome barriers to receiving vaccines, such as lack of transportation or the possibility of using the incentives for personal use among low socioeconomic individuals.

Thirdly, Technology-based health literacy is a method that can improve the overall acceptance of COVID-19 vaccines in China. This includes leveraging health literacy by using technologies such as posters, leaflets, social media platforms, and educational videos [67]. However, while doing so, it is important to ensure the creation of innovative educational information pieces that are engaging for the public. This will greatly maximize the engagement of the public and improve the knowledge regarding vaccines to address rumours, misconceptions, and concerns.

Additionally, interventions such as sending reminders through calls, text messages, and emails can function as media-based strategies to address vaccine hesitancy. These recall messages would help remind those individuals in China that are accepting COVID-19 vaccines and have not received the vaccine yet, possibly due to forgetting to book an appointment.

Overall, whilst working to pursue these educational-based approaches to address the current skepticism about vaccines, it is crucial to be mindful of jargon. While engaging with the public, layperson terminology should be used to convey scientific findings about the safety and efficacy of vaccines. This approach would help foster trust and a sense of belonging between the public and the scientific community and ultimately help boost the overall acceptance and uptake of COVID-19 vaccination services in China.

## 5. Conclusion

The results of this systematic review and meta-analysis map out the overall attitude of the population in China toward COVID-19 vaccines. An outcome faced following the COVID-19 pandemic and the creation of vaccines is vaccine resistance and hesitancy. This study showed that there is a notable variation with regard to vaccine acceptance and uptake in China across different population groups. There remains a deep-seated unwillingness and skepticism of the efficacy of COVID-19 vaccines amongst certain populations, such as patients with chronic diseases in China. Furthermore, this analysis uncovered statistically significant differences in the likelihood of vaccine uptake across various socio-demographic factors. Specifically, Chinese males and individuals with more than or equal to 5000 RMB had a higher COVID-19 vaccine uptake rate.

More studies are recommended to assess the behaviours of other population groups, such as remote employees, ethnic minorities, and religious people, to develop a broader understanding. Such studies would help evaluate the prevalence of vaccine uncertainty across diverse groups to help guide strategies to boost overall vaccine uptake. Vaccine hesitancy can prove to be a source of hindrance to vaccination campaigns and lessen the negative impacts of the COVID-19 pandemic. Furthermore, hesitancy regarding COVID-19 vaccines can also lead to the refusal of other routine immunizations. The prevalence of low willingness/acceptance and uptake of the COVID-19 vaccine mandates collaboration from the government, policy-makers, and media to campaign efforts to mitigate current barriers. It is recommended to focus on building trust between the public and government to prioritize clear communication and advocate for the need for vaccinations to achieve herd immunity among the overall population.

Future studies should also take into account key factors such as education level, residency status (rural/urban), and race to tailor current educational and vaccination programs according to the needs of the population. Novel research should also investigate other factors that lead to distrust and concern regarding COVID-19 vaccines by gathering the subjective experiences of the public. Overall, this study informs key considerations for the development of integrated models and a community-based transparent approach to guide future research and efforts.

## Data Availability

All relevant data are within the manuscript and its Supporting Information files.

## Acknowledgments

I wish to express my sincere gratitude to Dr. Zhou Xing for advising me throughout this scholarly paper project and for their insightful and valuable comments on the manuscript that helped to improve its quality to a great degree. This research received no external funding. All data utilized within this systematic review and meta-analysis is available in this manuscript.

## Supporting Information Captions

**S1 Table. Quality assessment** [19-65]

**S2 Table. GRADE Assessment for Odds Ratio - Age**

**S3 Table. GRADE Assessment for Odds Ratio - Gender**

**S4 Table. GRADE Assessment for Odds Ratio - Income**

**S5 Table. GRADE Assessment for Odds Ratio – Education**

**S6 Table. PRISMA Checklist**

**S1 Fig. Odds Ratio – Age funnel plot**

**S2 Fig. Odds Ratio – Gender funnel plot**

## Notes

### Competing Interest Statement

The authors have declared no competing interest.

### Funding Statement

The funders had no role in study design, data collection and analysis, decision to publish, or preparation of the manuscript.

